# Textual Triage: Assessing GPT4 for Classification of Free-Text Medication-Related Messages for Hypertension Management

**DOI:** 10.1101/2024.09.23.24314207

**Authors:** Ashley Batugo, Sy Hwang, Anahita Davoudi, Thaibinh Luong, Natalie Lee, Danielle L Mowery

## Abstract

Patient-generated free-text messages are a well-recognized source of clinical burden and burnout for clinicians. Machine learning approaches such as Large Language Models (LLMs) may be applied to alleviate this burden by automatically triaging and classifying messages, but their performance in this domain has not been fully characterized. In this study, we analyzed the effectiveness of GPT4 for classifying patient and provider messages for hypertension management through prompt engineering, comparing its performance to an alternative unsupervised generative statistical approach. The results of this study suggest GPT is promising for classification of medical-related messages even with very few guiding examples.

## Introduction

Free-text message exchanges, such as those afforded by electronic health portal messaging systems, provide an opportunity for clinicians to extend longitudinal care beyond the walls of traditional face-to-face clinics. However, these free-text messages are also an increasing source of clinical burden and burnout for clinicians^1-3^. Moreover, this burden is likely to grow as expanding digital health technologies further facilitate patient and provider communication. For example, many mobile health apps enable patient and provider communication through Short Messaging Service (SMS) for those with chronic conditions^4^. Solutions are urgently needed to improve care delivery experiences and outcomes for both patients and providers.

Natural Language Processing (NLP) and machine learning (ML) applications are promising potential tools to alleviate message burden. One potential application is to utilize NLP and ML approaches to triage patient messages to the appropriate member of the clinical team (scheduler, pharmacist, nurse, physician, etc.). There have been recent advances in the use of NLP and ML for filtering and reviewing clinical messages. In a study conducted by Chen et al. 2019, an NLP system called HypoDetect (Hypoglycemia Detector) automatically identified incidents of patient-reported hypoglycemia in secure message threads between patients with diabetes and the US Department of Veteran Affairs clinical teams^5^. Stenner et al. 2012 developed a rule-based NLP system called PASTE (Patient-Centered Automated SMS Tagging Engine) for extracting and tagging medication information from patient messages in a medication management system^6^.

Despite these promising examples, there are also substantial limitations to them. For HypoDetect, 3000 messages needed to be annotated for training and testing. For PASTE, the team only used existing libraries (RxNorm, RxTerms and NDF-RT); therefore, excluding the ability to subcategorize messages beyond what can be identified through these knowledge sources. In a previous related study conducted by Davoudi et al. 2022, investigators leveraged latent Dirichlet allocation (LDA), an unsupervised, generative statistical model for subgrouping observations in a dataset to see how well the LDA model could identify different medication related intent (goal or main idea of the text) categories^7^. While their results were promising, there was still much heterogeneity in intent for each LDA topic class. Even after applying a majority intent class heuristic (i.e. analysis was limited to messages containing only a single intent, while messages containing multiple intent categories were excluded), precision and recall values varied and some messages were not able to be predicted and classified.

ChatGPT, developed by OpenAI (San Francisco, CA, USA), is a large language model (LLM) trained on a large corpus of datasets using the generative pre-trained (GPT) architecture which utilizes neural networks to process natural language^8^. It can be leveraged to handle a wide variety of tasks including writing and debugging code^9,10^, answering exam questions from the United States Medical Licensing Examination (USMLE) Step 1 and Step 2 Exams at the level of a 3rd year medical student^11^, and diagnosing and triaging medical cases^12^. Because LLMs are already trained on a large corpus of data, and the knowledge acquired can be used for other downstream tasks, we hypothesized that its performance in message triaging and intent classification would be an improvement from our previous LDA approach. We also anticipated that it would have the advantage of requiring far less manual preparation and therefore sought to further discern its performance when provided zero, one, or three training examples (i.e. zero-, one-, or few-shot learning).

Thus, the objective of this study was to assess the performance of GPT4 for message classification. We assessed its performance on a set of medication related patient and provider messages for hypertension management, as previously described in Davoudi et al., 2022^7^. Briefly, these were text messages exchanged between patients and the health care team (nurse and physician) in an SMS-based remote hypertension management program. Messages for the study were manually reviewed, selected, and coded to create a dataset of messages that were limited to medication-related intent categories. The hypotheses of this study are as follows: H1) GPT4 can be highly accurate in classifying messages with recall and precision of above 0.9 and H2) Messages are best classified with few-shot learning.

## Methods

This study was reviewed and approved by the University of Pennsylvania Institute Review Board.

### Biomedical Data Overview

For this study, we obtained the de-identified and validated messages from the Davoudi et al study dataset^7^. This dataset included messages exchanged between providers and patients enrolled in Penn Medicine’s Employee Hypertension Management Program (eHTN) between June 2015 and November 2019. Through the program, participants were diagnosed with hypertension, received a prescription medication, and treatment plan for blood pressure (BP) management, and a BP cuff for conducting home-based readings. An essential component to this study was unlimited text message conversations between patients and providers for hypertension management through a proprietary Health Insurance Portability and Accountability Compliant (HIPAA) text messaging mobile app through Way to Health (W2H)^13^. The entire study consisted of messages exchanged between 253 participants and 5 providers (n=271 patient messages and 240 provider messages).

### Medication-Related Messages

The W2H dataset consists of short messages with one medication related intent that were manually annotated by two research team members.

### GPT4

We used GPT4 (temperature of 0.1) serviced as a private instance within Penn Medicine’s Microsoft Databricks tenant using the Azure OpenAI Service. When interacting with GPT4, data is not retained, and in this way, prompts are also not shared with the open source ChatGPT version. Additionally, because medical data is considered sensitive, Penn Medicine has opted out of OpenAI’s content filtering and management policy so that all input prompts and output responses are not be flagged. We interacted with GPT4 using the API and using the openai Python Package to build a custom conversational experience with GPT4, and to programmatically access GPT4. We classified messages using the GPT4 API (model: gpt4-32-k).

### Study Design

The study workflow (applies to both patient and provider messages) can be found in **Figure 1**. The complete dataset was separated into training and testing sets for both patient and provider messages. We used this strategy to assess whether our prompts would generalize well to unseen data. To avoid biased performance metrics, the final data set was split into 70% training and 30% testing data for both patient and provider messages. Using the training set, prompts were created for three experiments: 1) Experiment 1: zero-shot learning (providing no training examples), 2) Experiment 2: one-shot learning (providing one training example from the training set), 3) Experiment 3: few-shot learning (providing three training examples from the training set).

**Figure 1.**
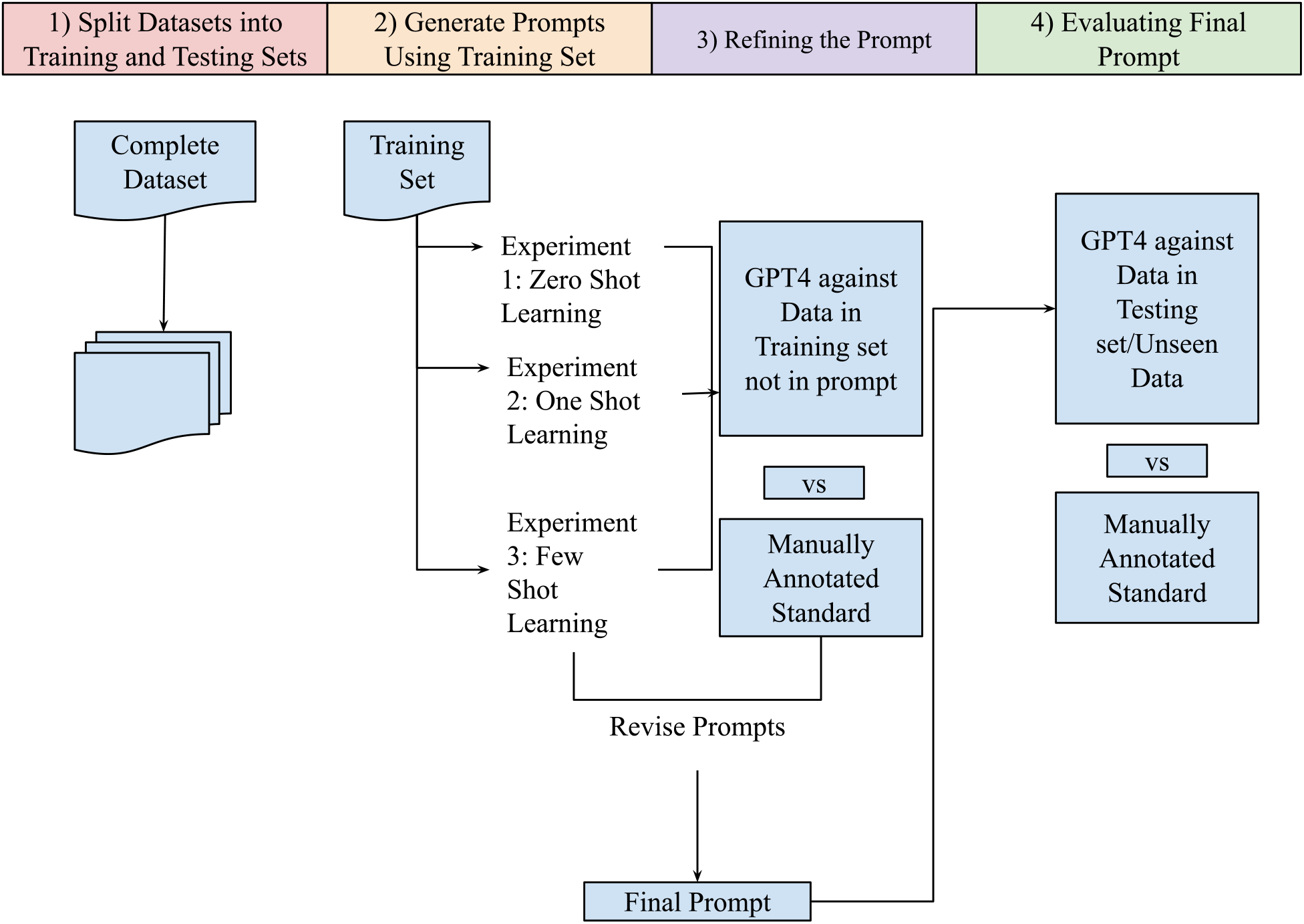
Study Workflow.

Additionally, the prompts include a role assigned to GPT4 (e.g. chatbot, triaging provider, etc.), an instruction prompt (the task it needs to do), descriptions of the message classes which were curated by manually reviewing the training sets, and sample messages and output for one and few shot learning. Below is an example prompt for patient message classification:

> *“You are a chatbot triaging messages from a text messaging system for patients experiencing hypertension with the goal to identify the intent of these messages so that they can be tagged and triaged to the appropriate healthcare provider. Your task is to conduct a precise binary classification for each message that comes through the chatbot system. Some messages may seem like they belong in multiple categories, but based on the descriptions for each of the message categories, assign labels as positive (1*.*0), negative (0*.*0) for belonging most to the target category. Below are the descriptions of each of the four classes of messages (1) medication_location, (2) medication_question, (3) medication_request, and (4) medication_taking: <description of each category>. This is the target category:<category name>. The following is a/are sample messages(s): <sample message(s)>. The following is/are the expected output(s): <1*.*0 or 0*.*0>”*.

After creating the prompts, they were sent to GPT4 for refinement. For each experiment, GPT4 was tasked to classify the rest of the training set as belonging to the specified medication intent category (1 for positive and 0 for negative) for the specified run in the prompt. GPT4s response was compared against the manually annotated reference standard. Until acceptable values for the performance metrics (precision and recall) was achieved, the prompts were continuously revised. The final prompts were then sent to GPT4 to classify the testing set. The results were then compared against the reference standard.

## Results

The training set used for curating prompts consisted of 195 patient messages and 170 provider messages. After dropping duplicates and because of issues with OpenAI Content Management Filtering Policy (though Penn Medicine has opted out of this^)14,^ the remaining training set consisted of 186 patient messages and 166 provider messages. The characteristics of this training set can be found in **Table 1**.

**Table 1.** Distribution of medication intent messages for training & testing set.

| Message Type<br>and Medication Intent Category | Messages, n (%) | Messages, n (%) |
| --- | --- | --- |
| Patient | Training (n=186) | Testing (n=85) |
| medication_taking | 52 (28.0) | 23 (26.8) |
| medication_request | 89 (47.8) | 41 (47.6) |
| medication_location | 36 (19.4) | 16 (19.5) |
| medication_question | 9 (4.8) | 5 (6.1) |
| Provider | Training (n=166) | Testing (n=74) |
| medication_refill | 2 (1.2) | 1 (1.4) |
| medication_change | 11 (6.6) | 5 (6.8) |
| medication_question_response | 25 (15.1) | 11 (15.1) |
| medication_question | 117 (70.5) | 52 (69.9) |
| medication_refill_question | 11 (6.6) | 5 (6.8) |

The training set messages not included in the prompts, which in this case were akin to a validation set, were used to tune the prompts. For zero-shot learning, all messages were used for evaluating GPT4’s performance, and for one-shot and few-shot learning, one message and three messages respectively, were excluded for evaluation. To measure the performance of GPT4 on the testing set in terms of precision defined as (true positive)/ (true positive + false positive) and recall defined as (true positive) / (true positive + false negative), it was tasked to classify 85 patient messages and 74 provider messages. Because of the content filtering issue, GPT4 was only able to classify 82 patient messages in the testing set. GPT4’s accuracy was very high for both patient and provider messages across all experiments. More than 89% and 90% of patients and provider messages, respectively, were accurately classified for all categories and across all experiments. The precision and recall metrics can be found in **Table 2**. For patient messages, GPT4 was able to identify all (Recall: 1.0) medication location and medication question messages for all experiments even though they only make up 19.51% and 6.10% of the data, respectively. Conversely, medication location messages had the overall highest precision and recall for the patient data. For provider messages, GPT4 was able to identify all (Recall: 1.0) medication refill question messages, despite only making up 6.85% of the training set. However, GPT4 had a precision and recall of both 0.00 for medication refill messages (which only consisted of one message in the training set). Additionally, every positive prediction made by GPT4 for patient medication request messages was correct (Precision: 1.0) for one and few-shot learning. Provider medication questions were the best classified across all experiments. GPT4s precision was lowest for zero-shot learning for the patient messages, and for provider messages, one-shot learning overall was the best approach. While GPT4’s performance was high to moderate for both precision and recall across all medication intents for patient messages, for providers messages, its performance was low-to-high for precision and recall across all medication intents. This might be because the provider testing set is much more imbalanced than the patient testing set.

**Table 2.** Performance of medication intent classification for testing set using n-shot learning.

| Message Type and Medication Intent Category | 0-Shot Learning |  | 1-Shot Learning |  | Few-Shot Learning |  |
| --- | --- | --- | --- | --- | --- | --- |
| *Patient* | *Precision (P)* | *Recall (R)* | *Precision (P)* | *Recall (R)* | *Precision (P)* | *Recall (R)* |
| medication taking | 0.74 | 0.91 | 0.83 | 0.91 | 0.87 | 0.91 |
| medication_request | 0.95 | 0.92 | 1.00 | 0.90 | 1.00 | 0.85 |
| medication_location | 0.73 | 1.00 | 0.76 | 1.00 | 0.89 | 1.00 |
| medication_question | 0.63 | 1.00 | 0.83 | 1.00 | 0.83 | 1.00 |
| Message Type and Medication Intent Category | 0-Shot Learning |  | 1-Shot Learning |  | Few-Shot Learning |  |
| *Provider* | *Precision (P)* | *Recall (R)* | *Precision (P)* | *Recall (R)* | *Precision (P)* | *Recall (R)* |
| medication_refill | 0.00 | 0.0 | NA | NA | NA | NA |
| medication_change | 0.50 | 1.00 | 0.71 | 1.00 | 0.67 | 0.80 |
| medication_question response | 0.67 | 0.55 | 1.00 | 0.73 | 0.86 | 0.55 |
| medication question | 0.91 | 0.96 | 0.92 | 0.92 | 0.92 | 0.92 |
| medication_refill_question | 0.41 | 1.00 | 0.56 | 1.00 | 0.56 | 1.00 |

In **Figures 2** and **3**, we show the outcomes of GPT4 message classification as belonging to a particular class (true positive (TP) and false positive (FP) predictions). The color of the bars indicates the manually annotated reference standard medication intent. The bars with a black outline are the TP results. Though GPT4 incorrectly classified many patient messages, many of these misclassifications were reduced after conducting few-shot learning. For the medication request messages, GPT4 incorrectly classified some medication location messages (n=2) as medication request messages, however those messages were no longer misclassified after one and few-shot learning.

**Figure 2.**
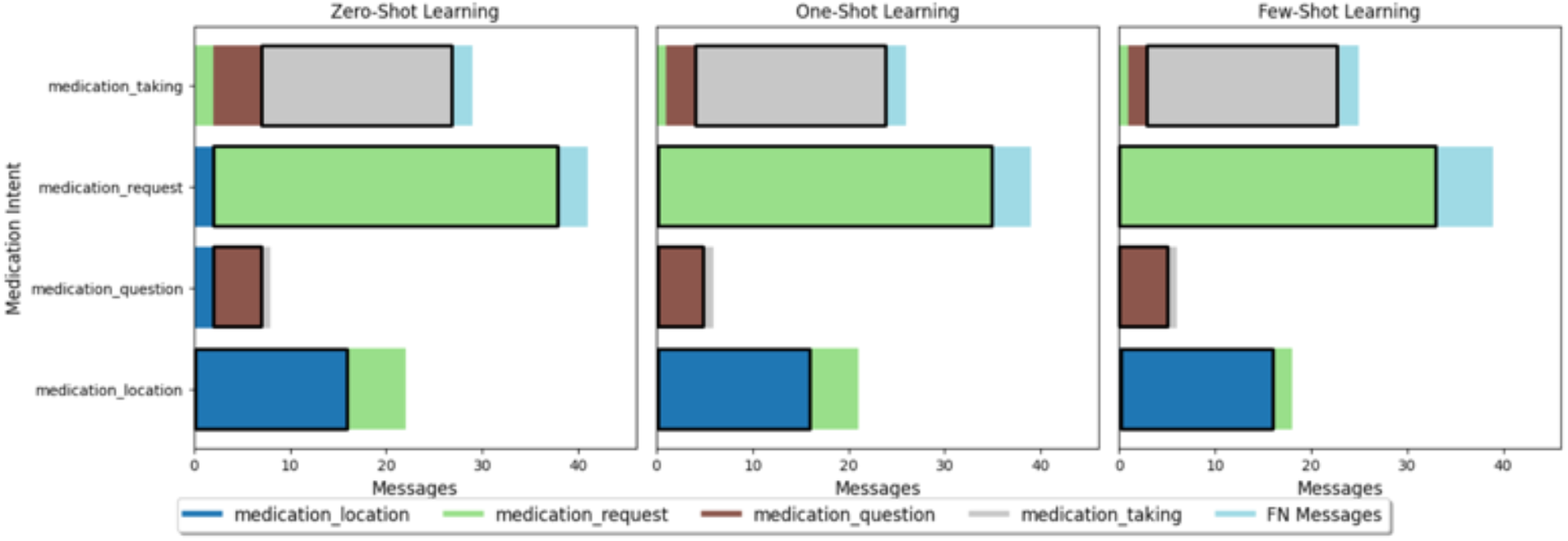
Distribution of medication intents for *patient messages* as classified by GPT.

**Figure 3.**
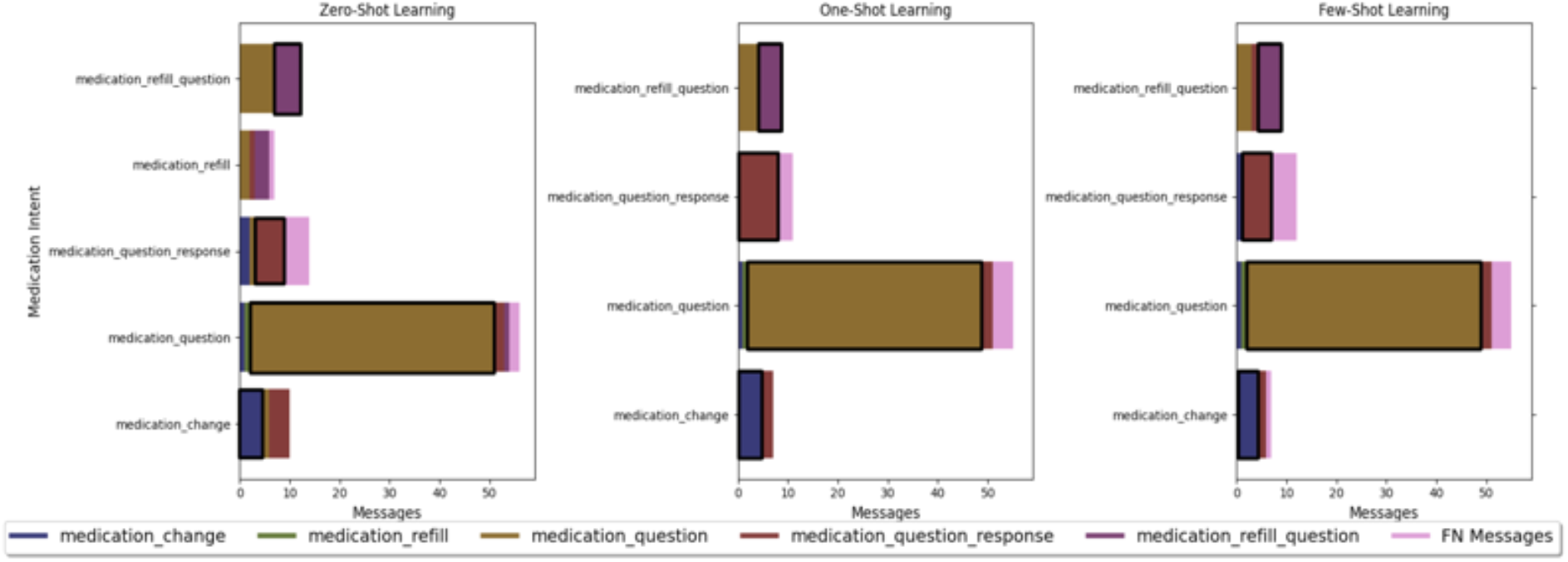
Distribution of medication intents for *provider messages* as classified.

For provider messages, GPT4 incorrectly classified messages for all categories across all experiments except for medication question response messages with one-shot learning. Additionally, GPT4 classified many messages that were medication questions as medication refill questions. However, this was decreased during one and few-shot learning. Generally, one and few-shot learning performed the same with one shot learning having a slightly better distribution for excluding FP messages in the provider set.

## Discussion

This study examined the performance of GPT4 to classify patient and provider messages exchanged in a bi-directional HIPAA-compliant text messaging mobile app. We found that GPT4 was able to classify patient messages with moderate to high precision and recall; however, because of the skewed data, it performed much worse on the provider set with low to high precision and recall. Because of this, GPT was not able to classify messages with precision and recall values of above 0.9. Compared to Davoudi et al.’s study^7^, GPT4 was able to correctly classify the messages in most cases better than, or in some cases comparable to LDA.

There are several important implications. First, GPT4 may be a promising tool for triaging free-text messages with reasonable performance. We used a fairly small (<300 messages total) and skewed training set for our experiments and still achieved good precision and recall; we anticipate that in contexts where GPT4 may actually be used for message classification, e.g. patient portal message triaging, this limitation might be overcome given the sheer volume of patient messages available. However, this study also suggests that GPT4 triage performance could suffer where the event of interest is infrequent (i.e. data are skewed).

Moreover, utilizing GPT4 for triage may require fairly minimal tailoring by the end-user. In our experiments, we found that predictive performance could be improved with just three additional training examples (few-shot learning). This makes GPT4 a flexible tool that can be adapted quickly to a specific local context and/or to dynamic clinical workflows, which are constantly being modified to adapt to health system needs. This is contrast to our primary comparator, LDA, or other machine learning approaches that often require large training sets to optimize performance.

This study is important because while LLMs represent a powerful new technology, its applications and roles within health care are still in development. One application under exploration is the use of LLMs to help respond to patient generated messages, but early on their impact is mixed. The majority of draft replies (80% or more, depending on clinician type) started by GPT4 were not used at all by clinicians in one pilot^15^, and in another study, common measures of EHR usage such as the amount of time required for clinicians to read or draft a reply, were not affected by the use of GPT4 responses^16^. This study suggests that another potential role for GPT4 is for message triaging.

This study also has some limitations and drawbacks. First, our dataset was limited to messages with a single intent category, which is not typical for free-text messages that often have layered, multiple intents. Further work is needed to assess GPT4 triage capability for more complex messages. Also, as mentioned above, we used a static Azure OpenAI GPT4 endpoint served on the Penn Medicine’s Databricks Platform. Despite choosing to opt-out of content filtering, many messages were excluded from being classified.

## Conclusions

This study found that GPT4 can classify medication related hypertension management messages exchanged between patients and providers. This model could also be used to classify other messages from this study with single intents and may even be used to improve the processes in which providers triage messages that come in through a patient’s portal. Consequently, this could provide patients with more timely care. Additionally, this provides an opportunity for extending communication between patients and providers beyond the patient portal as LLMs can help clinicians triage through messages that come by way of non-traditional means.

## Data Availability

Although this data has been de-identified prior to analysis, the data are not available.

## Acknowledgement

This project was supported, in part, by a grant from the National Institute on Aging (grant P30-AG034546), which provided financial support for AD and TL. NSL was funded by the Department of Veterans Affairs through the National Clinician Scholars Program. DM received funding from the National Institutes of Health for this work (grant UL1-TR001878). We also want to extend our gratitude to the Penn Data and Analytics Center of Excellence for administering the secure Microsoft Azure Databricks environment for this study.

